# Understanding depression in female adolescents: Correlations between interpersonal trust, self-esteem, and interpersonal attributional style

**DOI:** 10.1101/2023.02.09.23285684

**Authors:** Lin Li, Lan Tang, Yan-hong Lai, Jian Liu

## Abstract

This study examines sociocognitive factors causing depressed mood in hospitalized female adolescents with depression and the correlations between interpersonal trust, self-esteem, and interpersonal attributional style. From June 2021 to October 2022, a total of 166 hospitalized female adolescents with depression who met the inclusion criteria were recruited from Hangzhou Seventh People’s Hospital. They provided demographic information and were evaluated for depressed mood, interpersonal trust, self-esteem, and interpersonal attributional style using the Chinese versions of the Children’s Depression Inventory, Interpersonal Trust Scale, Rosenberg Self-Esteem Scale, and Multidimensional–Multiattributional Causality Scale, respectively. Interpersonal trust and self-esteem were negatively correlated with depressed mood, whereas internal and external attributions of interpersonal attributional style were significantly and positively correlated with depressed mood. Interpersonal trust was significantly and positively correlated with self-esteem, and interpersonal trust and self-esteem were negatively correlated with interpersonal attributional style. An analysis of the theoretical model for interpersonal trust–self-esteem– interpersonal attributional style–depressed mood relationships revealed that the model had a good fit and the overall fit indicators met the statistical requirements. The results of the path coefficient analysis indicated that all path coefficients reached statistically significant levels, except for those from interpersonal trust to depressed mood. Therefore, self-esteem mediates the effects of interpersonal trust and interpersonal attributional style on depression and is a critical sociocognitive factor affecting female adolescents with depression. Accordingly, psychological interventions for female adolescents with depression should focus on improving self-esteem.

## Introduction

Depression is a leading cause of adolescent illnesses and disabilities. International epidemiological studies report that the prevalence of adolescent depression is 1.8–7.8%, with a lifetime prevalence close to that of adults and a recurrence rate as high as 40–70% [1-3]. The latest epidemiological data from China indicate that the time-point prevalence of depressive disorder in children and adolescents aged 6–16 years is 3.0%, of which severe depression accounts for 2.0% [4]. Depressed mood is even more common. According to a recent report, the detection rate of various levels of depressive symptoms in adolescents has reached 24.6%; the risks of mild and severe depression are 17.2% and 7.4%, respectively. Notably, the detection rate of depressive symptoms in females is twice that in males [5]. This is why the present study focuses on female adolescents with depression (FADs). The onset of depression negatively affects adolescents’ social functioning and academic achievement and increases their risk of smoking, substance abuse, obesity, self-injury, and suicide [6]. In view of the high detection rates and negative impacts of depressive symptoms, the World Health Organization indicated that the primary disease burden in people aged 10–24 is major depressive disorder [7].

Interpersonal relationships are an important factor in adolescent depression [8,9]. According to evolutionary psychology, depression is an individual’s response to interpersonal pressure. When individuals fail to integrate into groups to meet their need for belongingness, they tend to exhibit signs of depression [10]. This view is unanimously supported by Chinese and international studies [11]. A Chinese report also indicated that the level of depression in adolescents increases proportionately with being unpopular among classmates, bullied, and criticized by teachers [5]. Conversely, protective factors against adolescent depression include high-quality peer relationships, positive parental educational methods, and positive teacher– student relationships [12,13]. Female adolescents are more likely to be affected by the quality of interpersonal relationships than male adolescents, because they focus more on the level of intimacy and loyalty in interpersonal relationships and are more likely to experience depressive reactions when such relationships break down [14].

Interpersonal relationships are affected by many sociocognitive factors, one of which is interpersonal trust (IT), an important topic in sociopsychological research. IT is a generalized psychological expectation established by individuals while interacting with others to determine whether their words and commitments (written or verbal) are honest and reliable [15]. Mutual trust is developed in the process of interpersonal communication and is based on the other party’s abilities and ethical standards [16]. As a link connecting individuals to other people and social development in general, IT is key to building harmonious relationships and social capital. It facilitates socioeconomic functioning, improves the operational efficiency of social organizations, and promotes economic growth [17].

Various studies have examined the relationship between IT and mental health in adolescents. IT has been reported to be critical to mental health development and an important facilitating force in forming and maintaining healthy interpersonal relationships [18,19]. Adolescents with high IT levels can better expand their social networks, thereby allowing them to gain higher social status, reduce loneliness, and establish high-quality peer relationships [20]. By contrast, the critical communication style that adolescents with depression adopt during their peer interactions generates negative emotions, which further exacerbates their feelings of rejection and isolation [21]. However, other researchers have suggested that IT does not predict the emergence of depression [22]. Some studies with Asian populations also found no correlation between IT and mental health issues [23]. Therefore, the impact of IT on depressed mood merits further investigation.

Self-esteem is another sociocognitive factor that affects interpersonal relationships. It is an overall evaluation of oneself and the positive attitude and high regard one has for all aspects of oneself [24]. The relationship between self-esteem and depressed mood has received extensive research attention. The susceptibility model of depression proposes that negative self-perception is a precursor to the development of depression in individuals and can predict its emergence [25]. This explanation is supported by several empirical studies, particularly those on late adolescents and adults. These studies consistently found that adolescents with low self-esteem were at a greater risk of and more likely to experience depression [26].

According to Erickson, the main task in adolescence is to develop one’s self-identity [14]. Hence, a consistent self-concept is crucial, as it determines whether individuals can transition to adulthood healthily and smoothly [14]. Adolescents acquire consistent information about themselves through diverse interpersonal interactions. The more affirmed they feel, the more positive their self-perception and the higher their self-esteem, which in turn affects the quality of their interpersonal relationships and the stability of their social support systems. When individuals enter adolescence, their focus transitions from their parents to peer groups to gain positive experiences. In this process, their self-esteem gains new dimensions for self-evaluation, namely, close friendships and attraction to those of the opposite gender [14].

Interpersonal attributional style (IAS), which similarly affects the establishment and maintenance of interpersonal relationships, is a cognitive process through which people draw conclusions about factors that influence or explain the consequences of their actions [24]. Various mental health effects arise from different attributional methods. The hopelessness theory of depression posits that certain features of people’s attributional methods are among the factors contributing to their depressed mood [27]. Specifically, individuals who tend to attribute bad events to themselves and view them as having long-lasting and comprehensive effects but attribute good events to others and view them as having temporary and localized effects are more likely to develop depressive symptoms [27]. This theoretical model has been supported by adequate evidence for the past 30 years. Initially intended for studying adult depression, some researchers have applied it to children and adolescents and verified that their negative attributional styles are related to depression [28].

The correlations between depression and the three important sociocognitive factors of IT, self-esteem, and IAS have been discussed to a certain extent. However, few studies have comprehensively examined the interactions among these three factors, which, in turn, affect the genesis or persistence of depression. Some studies have examined the impact of IT and self-esteem on mental health, while others have studied the impacts of self-esteem and attributional style instead. These studies concluded that self-esteem plays a mediating or moderating role in the relationship between depression and IT or attributional style [29-31]. Considering that IT, self-esteem, and IAS are important sociocognitive factors that contribute to stable interpersonal relationships and help adolescents obtain social support and maintain physical and mental health, we propose that examining the interactions between these factors can help clinicians better understand their role in predicting depressive symptoms.

Chinese and international studies have rarely sampled inpatient adolescents with depression to understand the mode by which IT, self-esteem, and IAS affect their depression. FADs are different from normal adolescents in terms of the causes and severity of depressed mood, resulting in more severely impaired social function, which has a greater negative social impact and leads to a higher disease burden. Relevant research findings can help patients, family members, teachers, and clinicians better understand the key sociocognitive factors affecting FADS; this can help identify the key points for intervention, improve the effectiveness of existing interventions, more effectively promote the recovery of social functions, and reduce the possibility of relapse, thereby reducing the societal disease burden. Therefore, hospitalized FADs were recruited to examine the mode by which IT, self-esteem, and IAS affect depression.

## Methods

### Participants

All data were acquired from a project titled “ Effects of interpersonal psychotherapy on the sociocognitive functions of adolescents with depression.” The research team recruited eligible FADs from hospitalized patients between June 2021 and October 2022. The inclusion criteria were (i) aged 12–18; (ii) diagnosed with depression according to the International Classification of Diseases 10th Revision diagnostic criteria and remained so upon discharge; (iii) relatively intact cognitive functions and capable of participating in a 1.5-hour-long interview and evaluation; (iv) no major physical disease, traumatic brain injury, or other organ damage; (v) no history of drug abuse or substance dependence; and (v) no concomitant diagnosis of other mental disorders.

There were 166 Han Chinese FADs, with an average age of 15.08±1.52 years. The general demographic data are presented in Table 1.

**Table 1.** General Demographic Data (N=166).

|  | Number | % |
| --- | --- | --- |
| *Age* | $15.08 \pm 1.52$ years | |
| *Grade of study* | $9.03 \pm 1.65$ | |
| *Place of residence* |  |  |
| City | 59 | 35.5 |
| Town | 71 | 42.8 |
| Village | 36 | 21.7 |
| *Only child* |  |  |
| Yes | 59 | 35.5 |
| No | 107 | 64.5 |
| *Father's occupation* |  |  |
| Public official | 13 | 7.8 |
| Company employee | 26 | 15.7 |
| Worker | 57 | 34.3 |
| Freelancer | 37 | 22.3 |
| Other | 33 | 19.9 |
| *Father's educational level* |  |  |
| Junior high school or below | 90 | 54.2 |
| High/vocational school | 40 | 24.1 |
| College or above | 36 | 21.7 |
| *Mother's occupation* |  |  |
| Public official | 17 | 10.2 |
| Company employee | 24 | 14.5 |
| Worker | 43 | 25.9 |
| Freelancer | 42 | 25.3 |
| Other | 40 | 24.1 |
| *Mother's educational level* |  |  |
| Junior high school or below | 98 | 59.0 |
| High/vocational school | 38 | 22.9 |
| College or above | 3 | 18.1 |
| *Annual household income (RMB)* |  |  |
| <100,000 | 62 | 37.3 |
| 100,000–300,000 | 71 | 42.8 |
| >300,000 | 33 | 19.9 |
| *Academic performance* |  |  |
| Top 5 | 6 | 3.6 |
| Above average | 44 | 26.5 |
| Average | 53 | 31.9 |
| Below average | 55 | 33.1 |
| Bottom 5 | 8 | 4.8 |
| *Duration of disease* | 20.62±18.28 months |  |
| *Number of hospitalizations* | 1.86±1.47 times |  |

### Evaluation indicators

#### Depressed mood

The Children’s Depression Inventory, compiled by Kovacs et al., is suitable for use with children aged 7–17 [32]. The questionnaire contains 27 items, each with three options describing the severity of depressive symptoms. The options are scored from 0 (least severe) to 2 (most severe) points. The total score is 54 points; the higher the score, the more severe the depression. The Chinese version of the scale has good reliability and validity among elementary and middle school children in China [32]. The Cronbach’s α coefficient in this study was 0.932.

#### IT

IT level was evaluated using Rotter’s Interpersonal Trust scale which comprises 25 items that include IT in various situations [33]. It can be used to predict an individual’s estimation of the reliability on other people’s behavioral commitments or statements. Scoring is based on a scale of 1 to 5; the higher the score, the greater the IT. The Chinese version of the scale has good reliability and validity [34], and the Cronbach’s α coefficient in this study was 0.826.

#### Self-esteem

The Rosenberg Self-Esteem Scale was used to evaluate adolescents’ overall feelings of self-worth and acceptance [35]. The 10 items are scored from 1 (completely not compliant) to 4 (completely compliant); the higher the score, the higher the self-esteem. The Chinese version of the scale has good reliability and validity [33], and its Cronbach’s α coefficient was 0.890 in the study.

#### IAS

The Multidimensional–Multiattributional Causality Scale, developed by Lefcourt et al., is divided into two subscales: academic achievement and IAS [36]. We used the Chinese version of the 24-item IAS subscale. The participants rated each item from 0 (completely disagree) to 4 (completely agree). The scale proposes four possible attributional categories: ability, effort, situation, and luck. According to the source of each attributional factor, ability and effort (situation and luck) are classified as internal (external) attribution. The scores for each dimension are calculated; the higher the score, the more likely the participants are to adopt the attributional method of the corresponding source. The Cronbach’s α for this subscale was 0.824.

### Research process

After the participants agreed to take part in the study and signed the informed consent, they would receive an evaluation on depressed mood, IT, self-esteem, and IAS. All of the questionnaires were self-reported. Partcipants were guided to the study room and to accomplish all of the evaluations assited by the graduate students trained in psychology, which lasted approximately 1.5 hours. Drug therapy, physical therapy, and other conventional treatments that the patients received during the study period, as well as the length of their hospital stay, were not affected by the study. All of the received data were recoded and the information which specify the identity of the participants were covered by new coding. The authors had no access to information that could identify individual participants after data collection.

### Statistical methods

Descriptive analysis was used to examine the patients’ general demographic data, which involved calculating the frequencies, means, and standard deviations of the variables. Additionally, the independent samples *t*-test, analysis of variance (ANOVA), and Pearson’s and Spearman’s correlation analyses were used to examine the impact of demographic variables on depressed mood. Next, descriptive analysis, the independent samples *t*-test, and ANOVA were used to analyze patients’ depressed mood, IT, self-esteem, and IAS. Pearson’s correlation analysis was also used to examine the relationship of depressed mood with IT, self-esteem, and the IAS.

The various statistical analyses were two-tailed tests and were performed using SPSS 25.0 (IBM Corp., Armonk, NY, USA). SPSS AMOS 24.0 (IBM Corp., Armonk, NY, USA) was used to test the relationship model for the effects of IT, self-esteem, and IAS on depressed mood. The maximum likelihood method was used to check the fit the model, and the fitting indicators included x^2^, degree of freedom (*df*), normed fit index (NFI), Tucker–Lewis index (TLI), goodness-of-fit index (GFI), comparative fit index (CFI), adjusted goodness-of-fit index (AGFI), root mean square error of approximation (RMSEA), and root mean square residual (RMR). The significance level for all statistical analyses was set to 0.05.

## Results

### Impact of demographic variables on depressed mood

The participants’ scores for depressed mood were subjected to an independent samples *t*-test or ANOVA under various categories of demographic variables (e.g., place of residence, being the only child, parents’ occupation and educational level, and annual household income). The results were not significant (*p*>0.05; Table 2).

**Table 2.** ANOVA Results for Depressed Mood Based on Demographic Variables.

|  | *F* or *t* value | *P* value |
| --- | --- | --- |
| Place of residence | 0.70 | 0.496 |
| Being the only child | 1.46 | 0.146 |
| Father's occupation | 0.66 | 0.654 |
| Father's educational level | 0.08 | 0.989 |
| Mother's occupation | 0.75 | 0.589 |
| Mother's educational level | 0.57 | 0.685 |
| Annual household income | 0.39 | 0.757 |
| Academic performance | 1.44 | 0.225 |

The results of the correlation analysis (Table 3) for age, grade of study, duration of disease, number of hospitalizations, and depressed mood revealed a negative correlation between age, grade of study, and depressed mood, which indicated that the older the participants and the higher their grade of study, the less often they experienced depressed mood. The number of hospitalizations and depressed mood were significantly and positively correlated; the greater the number of hospitalizations, the more severe was the depressed mood.

**Table 3.** Results of Correlation Analysis Between the Demographic Variables and Depressed Mood.

|  | *r* value | *P* value |
| --- | --- | --- |
| Age | -0.24 | 0.002 |
| Grade of study | -0.23 | 0.003 |
| Duration of disease | -0.01 | 0.953 |
| Number of hospitalizations | 0.17 | 0.031 |

### Descriptive outcomes of depressed mood, IT, self-esteem, and IAS

Table 4 presents the descriptive outcomes of the patients’ depressed mood, IT, self-esteem and IAS. The scores for depressed mood exceeded the threshold by 19 points, indicating an obvious depressed mood. The scores for IT were lower than the median score of 75 points. Patients’ external attribution (EA) tendencies were also more prominent. Significant differences were found in IAS scores across the four attributional categories: ability (2.42±0.71), effort (2.34±0.66), situation (2.79±0.59), and luck (2.27±0.66), *F*=20.818, *p*=0.000. The post hoc test indicated that the scores for the EA category of situation were the highest and significantly higher than those for luck, the other EA category, and the two internal attribution (IA) categories, ability and effort (*p*=0.000). Attribution to luck scored the lowest and was significantly lower than attribution to ability and the situation (*p*=0.035, *p*=0.00, respectively), although the difference in attribution to effort was not significant (*p*=0.350). There were no significant differences between the scores for the two IA categories of ability and effort (*p*=0.240).

**Table 4.** Descriptive Outcomes of Depressed Mood, IT, Self-Esteem, and IAS.

|  | *Average±SD* | *t* | *p* |
| --- | --- | --- | --- |
| Depressed mood | 26.78±11.71 |  |  |
| IT | 67.13±13.06 |  |  |
| Self-esteem | 23.17±6.54 |  |  |
| IAS |  |  |  |
| IA | 2.38±0.61 | -3.70 | 0.000 |
| EA | 2.53±0.54 |  |  |

Note: SD=standard deviation.

### Correlations between IT, self-esteem, IAS, and depressed mood

The correlations between IT, self-esteem, IA and EA, and depressed mood are shown in Table 5. Being in a depressed mood was negatively correlated with IT and self-esteem; the greater the IT, the higher the self-esteem and the lower the frequency of depressed mood. However, depressed mood was positively correlated with IA and EA; the higher the IA and EA scores, the more obvious the depressed mood.

**Table 5.** Correlations Between IT, Self-Esteem, IAS, and Depressed Mood.

|  | Depressed<br>mood | IT | Self-esteem | IA | EA |
| --- | --- | --- | --- | --- | --- |
| Depressed mood | 1 | -0.30** | -0.71** | 0.32** | 0.38** |
| IT |  | 1 | 0.31** | -0.20** | -0.19* |
| Self-esteem |  |  | 1 | -0.30** | -0.33** |
| IA |  |  |  | 1 | 0.59** |
| EA |  |  |  |  | 1 |

Note: \* $p < 0.05$ ; \*\* $p < 0.01$ .

There was a significant positive correlation between IT and self-esteem; the greater the IT, the higher the self-esteem. IT was significantly negatively correlated with IA and EA; the greater the IT, the lower the IA and EA scores. Self-esteem was significantly negatively correlated with IA and EA; the higher the self-esteem, the lower the IA and EA scores. A significant positive correlation was observed between IA and EA.

### Structural variance model for IT, self-esteem, IAS, and depressed mood

A theoretical model of the relationships between IT, self-esteem, IAS, and depressed mood was constructed based on existing research. The model was established after fitting existing data (x^2^=0.63, *df*=2, *p*=0.729). Table 6 presents the overall model fit indicators. The values of *x*^2^/*df*, NFI, TLI, CFI, GFI, and AGFI were above 0.95; RMSEA was less than 0.05, and RMR was 0.055.

**Table 6.** Overall Model Fit Indicators.

| Fit indicators | $\chi^2/df$ | NFI | TLI | CFI | GFI | AGFI | RMSEA | RMR |
| --- | --- | --- | --- | --- | --- | --- | --- | --- |
| Standard deviation | <2 | $\geq 0.95$ | $\geq 0.95$ | $\geq 0.95$ | $\geq 0.95$ | $\geq 0.95$ | $\leq 0.05$ | $\leq 0.05$ |
| Numerical value | 0.316 | 0.997 | 1.03 | 1.000 | 0.998 | 0.989 | 0.000 | 0.055 |

Hu and Bentler stated that the model fit is good when *x*^2^/*df* is 0–2; the values of NFI, TLI, GFI, AGFI, and CFI are above 0.95; and those of RMSEA and RMR are not higher than 0.05 [37]. The model is acceptable when *x*^2^/*df* is 2–5; the values of NFI, TLI, GFI, AGFI, and CFI are above 0.9; and those of RMSEA and RMR are less than 0.1. All data in this study met the statistical requirements, indicating good overall model fit.

The path coefficient analysis revealed that the path coefficients reached statistically significant levels (*p*<0.05), except for those from IT to depressed mood. The details of the relevant path coefficients and diagram of the model are presented in Table 7 and Fig 1, respectively.

**Table 7.** Significance Test for the Path Coefficients.

| Path | | | Standard<br>regression<br>coefficient ($\beta$) | SE | CR | P |
| --- | --- | --- | --- | --- | --- | --- |
| IAS | → | Self-esteem | -0.35 | 0.112 | -3.850 | 0.000 |
|  |  | Depressed<br>mood | 0.20 | 0.160 | 2.727 | 0.006 |
|  |  | IA | 0.72 |  |  |  |
|  |  | EA | 0.81 | 0.190 | 5.227 | 0.000 |
| IT | → | IAS | -0.25 | 0.038 | -2.670 | 0.008 |
|  |  | Self-esteem | 0.23 | 0.037 | 3.052 | 0.002 |
|  |  | Depressed<br>mood | -0.06 | 0.051 | -1.041 | 0.298 |
| Self-esteem | → | Depressed<br>mood | -0.61 | 0.112 | -9.688 | 0.000 |

**Fig 1.**
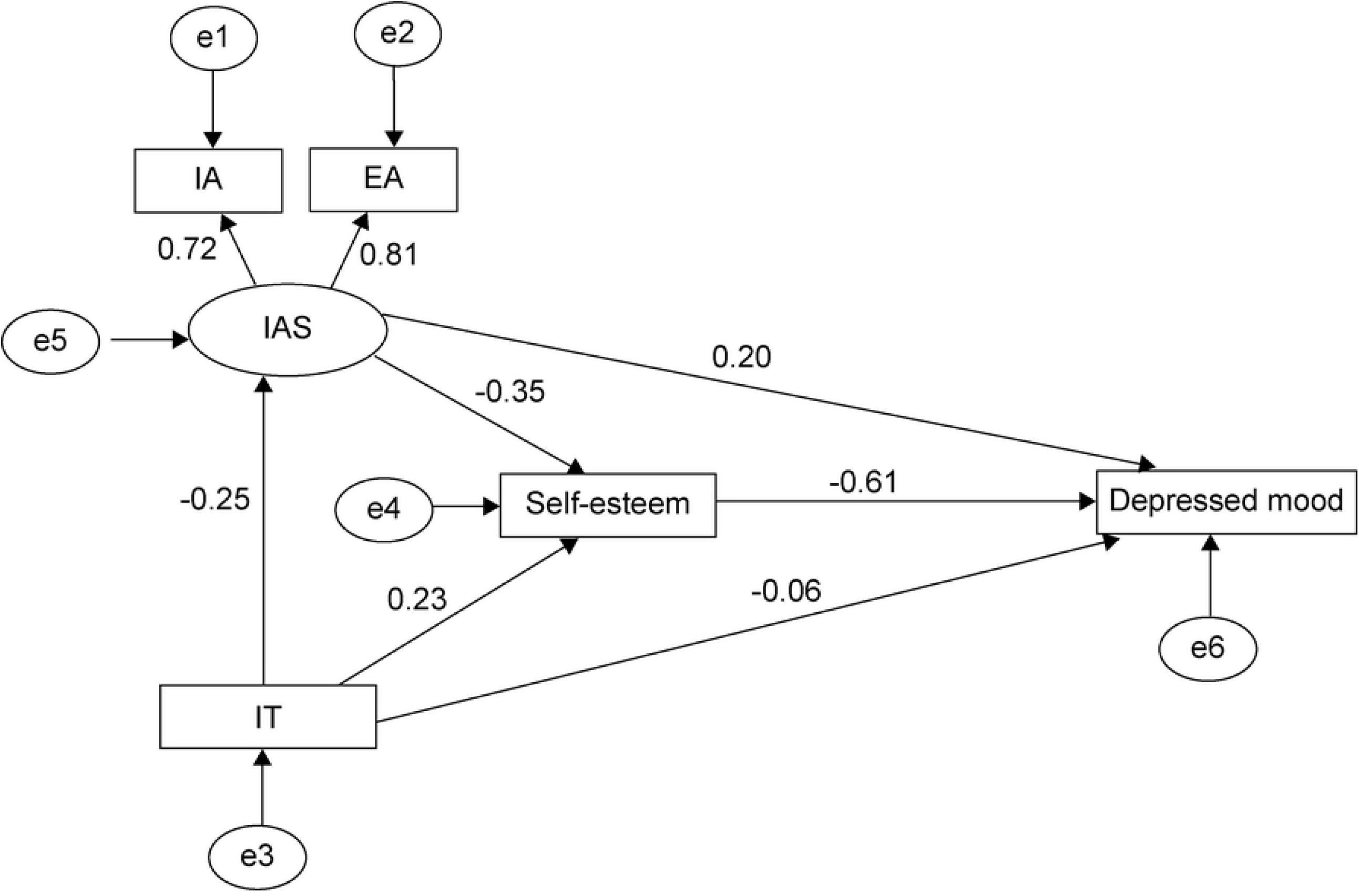
Model for the impact of IT, self-esteem, and IAS on depressed mood Note: IA: internal attribution; EA: external attribution; IAS: interpersonal attributional style; IT: interpersonal trust; e1-e6: error.

IT affected depressed mood through three paths (Fig 1). The first was through self-esteem, which had a complete mediating effect in the relationship between IT and depressed mood. The second was through the complete mediating effect of IAS. The dual mediating effects of IAS and self-esteem formed the third path.

IAS could affect depressed mood on its own or through the partial mediating effect of SE. As an important mediating variable in this model, self-esteem not only affected the relationship between IT and depressed mood but also moderated the effect of IAS on depressed mood.

## Discussion

In this study, depressed mood, IT, self-esteem, and IAS of 166 hospitalized FADs were investigated and analyzed. The results showed that the patients had obvious depression when they were enrolled in the study, and their IT and self-esteem scores were average. For IAS, participants were more inclined toward the EA category. IT, self-esteem, and IAS (IA and EA) were significantly correlated with depressed mood. In the relationship model for the four variables, the relationship between IT and depressed mood was affected by two factors: SE and IAS. IT required the mediating effect of self-esteem or IAS to affect depressed mood. Although IAS could directly affect depressed mood, its effect was partially mediated by the introduction of self-esteem. As an important mediating variable, self-esteem not only moderated the effects of IT and IAS on depressed mood but also produced the most important predictive effect on depressed mood for the entire pathway.

There is no consistent conclusion regarding the effect of IT on adolescent depression. Adolescents with good IT fared better than those with poor IT in expanding their social networks, gaining high-quality peer relationships and high social status, and reducing loneliness [20]. However, this impact may be unstable [22,23,38]. Further, there may be other factors that exert mediating or moderating effects in the relationship between IT and depressed mood [20,39,40]. McCarthy et al. proposed that the personality traits of self-esteem and agreeableness could predict IT and the revelation of negative emotions [41]. Through diary studies, Li et al. established that self-esteem modulates the correlation between self-exposure and IT when forming offline friendships [42]. From their study of the relationship between social avoidance and depressed mood in 1,021 college students, Yuan et al. concluded that IT mediated the correlation between social avoidance and depressed mood, whereas self-esteem moderated the effect of social avoidance on IT [43].

The aforementioned studies focused on college students. By contrast, we surveyed hospitalized FADs, and the results indicated that self-esteem and IAS affected the correlation between IT and depressed mood. When negative events continued to occur in the FADs’ relationships and their IT was destroyed, their self-esteem and IAS determined whether they would experience depressed mood. The lower the self-esteem, the more obvious the application of EA, and the more likely the appearance of depressed mood. However, high self-esteem and flexible IAS can moderate the negative emotional effects caused by adverse interpersonal events and damaged IT.

There has been extensive research and consensus on the relationship between IAS and depressed mood [44]. Our findings indicated that IAS was significantly positively correlated with depressed mood in hospitalized FADs but significantly negatively correlated with self-esteem and IT. Self-esteem also mediated the effect of IAS on depressed mood. The patients who participated in this study were more inclined to develop EA in their IAS. Attributing interpersonal events to situations is externally controlled and more difficult for individuals to control with their will [24]. Individuals who do so are more likely to feel frustrated and lost during their interpersonal interactions. This conclusion supports the hopelessness/self-esteem model for depression. This model, proposed by Metalsky et al., integrates the susceptibility model of depression and self-esteem theory and proposes that negative attributional methods generate hopelessness and depressed mood through low self-esteem and failures. By contrast, high self-esteem plays a buffering role in the process of attribution failures that cause the development of depression [30].

The effect of self-esteem on adolescents’ emotional problems has been supported [25,26]. Adolescents with low self-esteem report more emotional and behavioral issues such as suicidal ideation, self-injury, depression, and anxiety [45]. The susceptibility model of depression posits that self-esteem is an important risk factor for the development of depression [25]. This model is supported by our findings: self-esteem was the most significant factor that affected depressed mood and could mediate the effects of IT and IAS on depressed mood. This is also in line with the characteristics of psychological development during adolescence. Adolescents pay more attention to their self-image and wish to portray a consistent image. Positive and stable self-evaluation determines an individual’s emotional responses when facing adverse life events. This is beneficial because individuals are protected and can stably weather the turbulent adolescent period. They can also complete the successful establishment of their self-identity and transition into healthy adulthood [14].

Additionally, we found that the grade of study, age, and number of hospitalizations affected the severity of depressed mood. The older the patient, the higher the grade, and the fewer the hospitalizations, the less often they experienced depressed mood; the younger the patient, the lower the grade, and the more the hospitalizations, the more frequent the depressed mood. This may be related to the extent of physical, cognitive, and social development in female adolescents. In females, the physiological changes associated with puberty generally begin and end around the age of 10 and 14 years, respectively. During this stage, imbalances in physical and psychological development are more obvious, and emotional fluctuations are more prominent. The incomplete development of cognitive functions is a limitation, and self-identity is yet to be established. Explosive expressions of female emotions occur more frequently during early adolescence. Parents easily perceive such obvious changes and become more concerned about the situation [5,14].

This study explored the sociocognitive factors affecting FADs and found that self-esteem is an important variable that regulates or mediates the relationships of IT and IAS with depressed mood. Based on our findings, the following are some novel suggestions for the psychological treatment of FADs. First, early intervention for FADs is critical, and the number of hospitalizations can be effectively reduced with early treatment, thereby alleviating the severity of their depressed mood and helping them navigate through adolescence more smoothly. Second, interpersonal relationships are an important theme in adolescents’ development and are related to their mental health status. The key to helping FADs rebuild trust in interpersonal communication and reduce depression during interpersonal psychotherapy is improving their self-esteem and reducing their tendency for EA in interpersonal relationships. This can, in turn, facilitate the establishment of trust in interpersonal communication, which leads to the development of high-quality interpersonal relationships, thereby allowing them to gain additional social support.

This study also has some limitations. First, it is a pilot study, and representative samples are still needed for further verification of the results. Second, as IT, self-esteem, and IAS are multidimensional concepts, the questionnaire adopted in this study cannot offer a comprehensive perspective. Moreover, the use of self-report questionnaires lead to a lack of ecological validity. The causal relationship between these real behavioral outcomes and the emergence of depression warrants further research and exploration.

## Data Availability

The datasets used and/or analysed during the current study are available from the corresponding author on reasonable request.

